# Frequency and phenotype associations of rare variants in five monogenic cerebral small vessel disease genes in 200,000 UK Biobank participants with whole exome sequencing data

**DOI:** 10.1101/2021.11.17.21266447

**Authors:** Amy C. Ferguson, Sophie Thrippleton, David E. Henshall, Ed Whittaker, Bryan Conway, Malcolm MacLeod, Rainer Malik, Konrad Rawlik, Albert Tenesa, Cathie Sudlow, Kristiina Rannikmäe

## Abstract

Based on previous case reports and disease-based cohorts, a minority of patients with cerebral small vessel disease (cSVD) have a monogenic cause, with many also manifesting extra-cerebral phenotypes. We investigated the frequency, penetrance, and phenotype associations of rare variants in cSVD genes in UK Biobank (UKB), a large population-based study.

We used a systematic review of previous literature and ClinVar to identify putative pathogenic rare variants in *CTSA, TREX1, HTRA1, COL4A1/2*. We mapped phenotypes previously attributed to these variants (phenotypes-of-interest) to disease coding systems used in UKB’s linked health data from UK hospital admissions, death records and primary care. Among 199,313 exome-sequenced UKB participants, we assessed: the proportion of participants carrying ≥1 variant(s); phenotype-of-interest penetrance; and the association between variant carrier status and phenotypes-of-interest using a binary (any phenotype present/absent) and phenotype burden (linear score of the number of phenotypes a participant possessed) approach.

Among UKB participants, 0.5% had ≥1 variant(s) in studied genes. Using hospital admission and death records, 4-20% of variant carriers per gene had an associated phenotype. This increased to 7-55% when including primary care records. Only *COL4A1* variant carrier-status was significantly associated with having ≥1 phenotype-of-interest and a higher phenotype score (OR=1.29, p=0.006).

While putative pathogenic rare variants in monogenic cSVD genes occur in 1:200 people in the UKB population, only around half of variant carriers have a relevant disease phenotype recorded in their linked health data. We could not replicate most previously reported gene-phenotype associations, suggesting lower penetrance rates, overestimated pathogenicity and/or limited statistical power.

## Introduction

Cerebral small vessel disease (cSVD) refers to a variety of pathological processes impacting the brain’s small arteries, arterioles, venules, and capillaries^1^. It is estimated that 20% of ischaemic strokes, and most haemorrhagic strokes, are caused by cSVD. cSVD is also the most frequent pathology underlying vascular dementia and vascular cognitive impairment^2–4^.

An unknown minority of cSVD cases are considered monogenic, i.e., caused by a pathogenic variant(s) in one of several genes. While *NOTCH3* (implicated in cerebral autosomal dominant arteriopathy with subcortical infarcts and leukoencephalopathy, CADASIL) is the best known of these, since its first description in 1996^5^, several additional cSVD genes have been identified. Examples include *CTSA, TREX1, HTRA1, COL4A1* and *COL4A2*, but there are additional genes where cSVD is either not the primary associated phenotype (e.g., *ADA2* and *GLA*) or to date there is weaker causal evidence (e.g., *FOXC1, PITX2, COLGALT1*)^2^. Many monogenic cSVD cases also show overlapping systemic and neurological features^2,6,7^. Our recent systematic literature review of ⍰800 individual cases carrying putative pathogenic variants in six cSVD genes found that several monogenic cSVD genes are associated with a range of extra-cerebral phenotypes affecting ocular, renal, hepatic, muscle, and haematological systems^6^. These extra-cerebral phenotypes can be diagnostically important through contributing to clinically recognisable syndromes informing genetic testing, as well as provide mechanistic insights into the pathophysiology of cSVD^6,8,9^.

To date, our knowledge of monogenic cSVD variants’ frequency, penetrance and phenotype associations comes primarily from case reports, small case series and family pedigree studies^6^. The resulting data is therefore affected by various biases, including investigation bias (patients with clinically severe and previously described manifestations are more likely to have genetic testing and undergo investigations for known expected associated pathologies if a pathogenic variant in a relevant gene is found), publication bias (clinicians/researchers are more likely to publish a case report/series about clinically severely and/or unusually affected patients), and reporting bias (published case reports/series tend to discuss previously reported or particularly unusual clinical signs and symptoms rather than describe case’s health in an unbiased and systematic way)^6,10,11^. There have also been few disease-based studies exploring rare variation in cSVD genes in apparently sporadic cases of cSVD^12–15^, but the population frequency and clinical consequences of these variants remains unknown.

Data from large-scale population-based studies collecting health outcomes in a systematic, unbiased way would provide additional information on the frequency and clinical impact of monogenic cSVD rare variants in a different setting. Investigating routinely collected, linked, healthcare records would overcome some of the limitations of previous studies.

The UK Biobank (UKB) cohort is a prospective study of approximately 500,000 UK residents recruited from 2006-2010, aged 40-69 at the time of recruitment. It has extensive phenotypic information derived from linked healthcare and death record data, with whole exome sequencing (WES) data available for around 200,000 participants as of October 2020. Full details of UKB have been previously described^16–18^.

We aimed to use UKB to assess the population frequency of putative pathogenic rare genetic variation in five known monogenic cSVD genes - *CTSA, TREX1, HTRA1, COL4A1* and *COL4A2* (excluding *NOTCH3* since it has already been investigated in UKB^19^). We studied their apparent penetrance (i.e., the proportion of participants with a variant manifesting a relevant cerebral and/or extra-cerebral clinical phenotype) and gene-phenotype associations in the general population, not selected on the basis of disease or disease risk (Figure 1).

**Figure 1.**
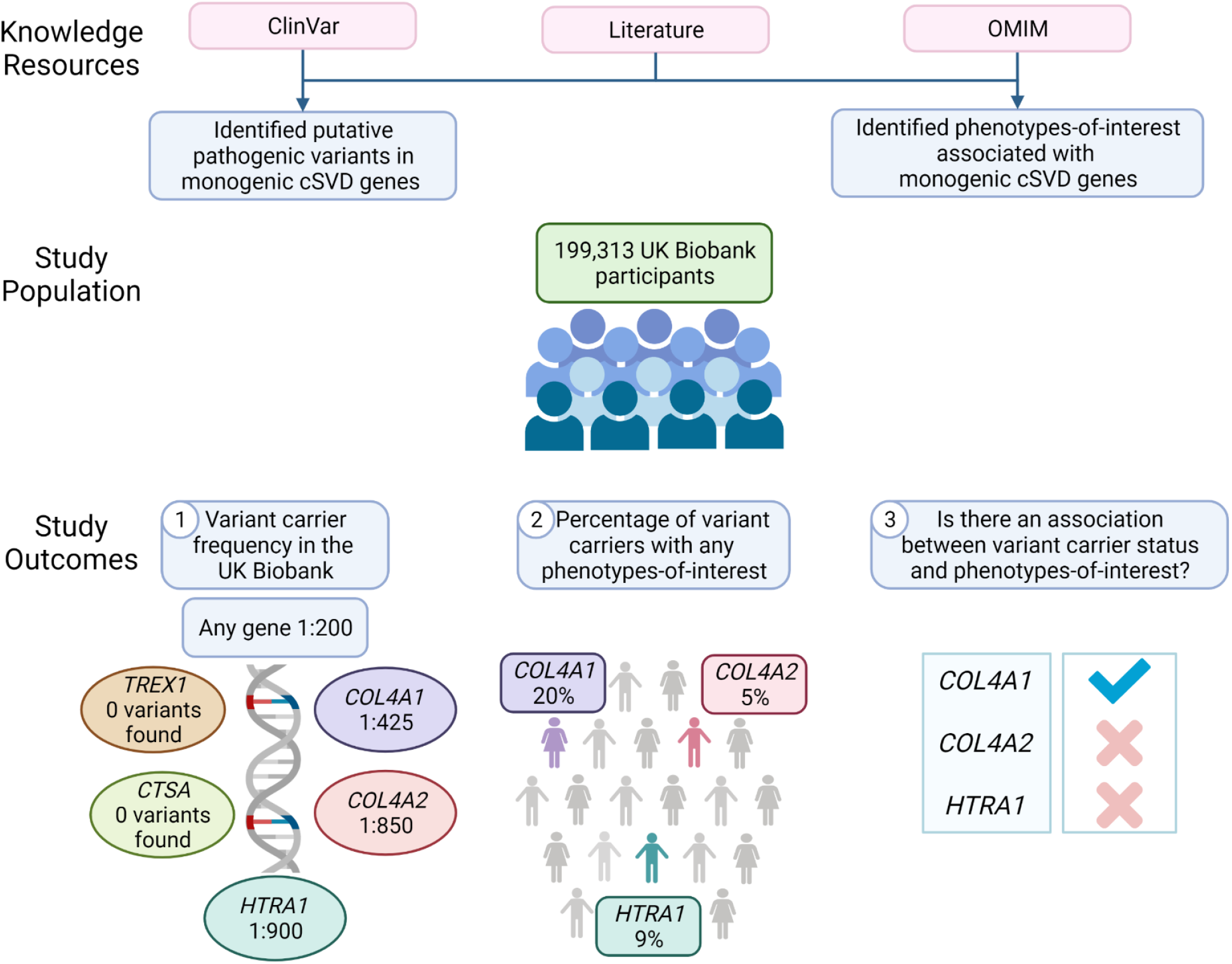
Summary of study methods and outcomes. OMIM=Online Mendelian Inheritance in Man; cSVD=cerebral small vessel disease. Created with BioRender.com

## Methods

### Study population

Our population of interest comprised the 200,603 UKB participants with WES data available from October 2020. We excluded: (i) related individuals based on genetic relatedness pairings as provided by UKB Field 22011, excluding one individual from each related pair but preferentially retaining participants carrying a variant-of-interest; and (ii) participants with sex mismatch (reported sex did not match genetic recorded sex). After these quality control exclusions, our study population comprised 199,313 UKB participants. The following data was available for the whole sample: (i) WES; (ii) coded hospital inpatient admissions and death record data [UKB Fields 41270, 40001, 40002]; (iii) baseline characteristics (age at last follow up on March 2020 [derived from UKB Fields 34 and 52], sex [UKB Fields 31 and 22001], self-reported ethnicity [UKB Field 21000], Townsend deprivation index – a marker of socio-economic deprivation [UKB Field 189]). Coded primary care data [UKB Field 42040] was also available for a 48% subset (95,459 participants).

### Variant selection

We identified putative pathogenic rare variants in *CTSA, TREX1, HTRA1, COL4A1* and *COL4A2*, which we refer to as ‘variants-of-interest’. We defined putative pathogenic rare variants as variants that are: (i) reported as causing disease in the published literature based on our systematic review (SysRev variants)^6^; and/or (ii) reported as “pathogenic” or “likely pathogenic” in the ClinVar database (ClinVar variants)^20^; and (iii) have a minor allele frequency (MAF) <1% in the UK Biobank. An exception to this were *TREX1* and *CTSA* genes, which in addition to monogenic cSVD are also associated with other specific monogenic disorders (Aicardi-Goutieres syndrome and Galactosialidosis, respectively) ^21–23^. Hence for these two genes, variants reported to be specific for conditions other than cSVD were excluded. We used the Ensembl variant effect predictor (VEP) to estimate the pathogenicity and protein impact of variants-of-interest, based on SIFT, PolyPhen and SnpEff^24^.

### Phenotype selection and mapping

We first compiled a list of cerebral and extra-cerebral phenotypes previously attributed to *CTSA, TREX1, HTRA1, COL4A1* and *COL4A2* variants-of-interest. We included phenotypes reported as being associated with these genes in Online Mendelian Inheritance in Man (OMIM) and/or in our recent systematic review^6,23^, from here-on referred to as phenotypes-of-interest. We mapped these phenotypes to the disease-coding systems used for recording hospital inpatient admissions, death record and primary care data in the UKB, i.e., International Classifications of Diseases – 10 (ICD-10), Read V2 and Read V3 disease-coding systems. Further detail regarding the mapping process is provided in the Supplemental Methods.

### Data analyses

#### Assessing variant carrier frequencies in the UKB and their demographic characteristics

Using Functional Equivalence (FE)-derived PLINK files^17,18^, we calculated the total number and proportion of UKB participants with ≥1 variant-of-interest (from here-on referred to as “variant carriers”). We used Chi-squared and two sample t-tests to assess differences between variant carriers and non-carriers in age, sex, Townsend deprivation index, ethnicity, and the presence of vascular risk factors (0-1 versus ≥2 risk factors described in Supplemental Table 6 legend).

#### Assessing the proportion of variant carriers with phenotypes-of-interest

As a measure of genetic variant penetrance, we calculated the proportion of variant carriers with ≥1 phenotype-of-interest in the hospital inpatient admissions and/or death record data for the whole study population, and in hospital admissions, death record and/or primary care data for the 48% subset of the study population with linked primary care data. We further explored the proportion of variant carriers with a stroke diagnosis, one of the main clinical manifestations of cSVD^2–4^.

#### Assessing whether variant carrier status is associated with phenotypes-of-interest

We tested for statistically significant associations between variant carrier status and phenotypes-of-interest by gene. We undertook the primary analyses in the whole cohort and repeated these as secondary analyses in the subgroup with primary care data.

### Association with phenotype-of-interest status

For each of the five genes, we checked for differences in the proportion of participants with any phenotype-of-interest, and with stroke specifically, among variant carriers compared to non-carriers. We used a Chi-squared test and set a Bonferroni-corrected significance p-value of <0.01 (corrected for the 5 gene-level tests).

### Association with phenotype burden (phenotype score)

We assessed for difference in overall phenotype burden between variant carriers and non-carriers, creating for each gene and participant: binary variant scores using an unweighted gene-based collapsing approach^25^ (with carriers given a score of 1 and non-carriers a score of 0); and unweighted phenotype scores (based on the number of the phenotypes-of-interest associated with the gene). For example, *HTRA1* is associated with 6 phenotypes-of-interest and a participant could therefore have a minimum *HTRA1* phenotype score of 0 (if they did not manifest any phenotypes-of-interest) and a maximum score of 6 (if they manifested all 6 phenotypes-of-interest) (Supplemental Table 1). We used Poisson regression (due to the phenotype scores being based on exact counts) to investigate, for each gene, the association between carrying a rare variant and phenotype score including age at last follow-up, sex, Townsend index, and 20 genetic principal components (PCs) as covariates. We used a Bonferroni-corrected p-value threshold of <0.01 to determine significance (corrected for 5 gene-level tests).

For primary analyses we additionally: (i) adjusted for vascular risk factors ^26,27^ (Supplemental Table 6 legend); (ii) checked for interactions between variant carrier status with sex and ethnicity; and (iii) ran leave-one-out sensitivity analyses, where each phenotype was removed from the phenotype score one-by-one (using logistic instead of Poisson regression in case of *COL4A2* which only had two phenotypes-of-interest to start with).

## Results

We identified a total 253 variants-of-interest across the five genes to investigate in UKB: 145 exclusively from SysRev, 48 exclusively from ClinVar, and 60 from both sources. We found a total of 37 variants present in ≥1 of the 199,313 included UKB participants: 24 exclusively from SysRev, 2 exclusively from ClinVar, and 11 from both sources, but these did not include any *TREX1* or *CTSA* variants-of-interest. The number of variants represented in UKB varied for each gene, ranging from 5 *(COL4A2)* to 17 *(COL4A1)*. Across these variants, MAF in UKB ranged from 0.0005-0.14%. (Supplemental tables 2 and 3).

VEP predicted 92% (22/24) SysRev, 100% (2/2) ClinVar and 100% (11/11) variants from both sources to be missense or nonsense mutations. Of the remaining two SysRev variants, one was in the 5’ untranslated region and the other an intronic splice donor variant. SnpEff predicted the nonsense mutations and the splice donor variant to have high impact, while all but one of the remaining variants were of moderate impact. Overall, 21/37 variants (50% SysRev; 50% ClinVar, 73% both sources) were predicted to have probably damaging and deleterious impacts on protein structure and function (Supplemental Table 4).

We identified 2 to 12 phenotypes-of-interest per gene (Supplemental table 1). When mapping these to disease coding systems, the number of codes per phenotype varied widely, ranging from 6 codes for dry mouth to 173 codes for degenerative spine disease (Supplemental code list).

### Assessing variant carrier frequencies in the UKB and their demographic characteristics

Among 199,313 UKB participants, 1,050 participants (0.5%) had ≥1 variant-of-interest, resulting in 234 *HTRA1*, 481 *COL4A1* and 336 *COL4A2* variant carriers, with one participant carrying a variant in both *HTRA1* and *COL4A1*. When variant carriers were not preferentially selected from related pairs, their overall frequency remained the same. Most variant carriers (96%; 1,003/1,050) possessed a SysRev variant, 0.3% (3/1,050) a ClinVar variant, and 4% (44/1,050) a variant represented in both SysRev and ClinVar (Supplemental tables 2 and 3).

The mean age (at last follow-up) of all included UKB participants was 68.2 years. The mean Townsend deprivation index was -1.34 and 55% of participants were female. There was no significant difference in age at last follow-up, sex, or presence of vascular risk factors between variant carriers and non-carriers. Carriers had significantly higher levels of deprivation than non-carriers (mean Townsend index = -0.44 vs -1.34, p<2.2×10^−16^), and were more likely to be of non-white ethnicity (p<2.2×10^−16^) (Table 1).

**Table 1.** Demographic characteristics of UK Biobank participants with WES data.

|  | All UKB participants with WES<br>(N=199,313) | Variant carriers<br>(N=1,050) | Non-carriers<br>(N=198,263) | p-value |
| --- | --- | --- | --- | --- |
| Age at last follow up<sup>1</sup> |  |  |  |  |
| Mean (SD) | 68.2 (0.018) | 67.7 (0.26) | 68.2 (0.018) | p=0.43 |
| Sex |  |  |  |  |
| Female | 109,739 (55%) | 589 (56%) | 109,150 (55%) | p=0.52 |
| Male | 89,574 (45%) | 461 (44%) | 89,113 (45%) |  |
| Townsend Deprivation Score |  |  |  |  |
| Mean (SD) | -1.34 (0.01) | -0.44 (0.11) | -1.34 (0.01) | p<2.2x10<sup>-16</sup> |
| Self-reported ethnic group<sup>2</sup> |  |  |  |  |
| White | 187,035 (94%) | 783 (75%) | 186,252 (94%) | p<2.2x10<sup>-16</sup> |
| Non-White | 11,319 (6%) | 256 (25%) | 11,063 (6%) |  |

Legend: N=number of participants; UKB=UK Biobank; WES=whole exome sequencing; SD=standard deviation; <sup>1</sup>Age at last follow up is the participant's age in March 2020, when the linked health record data is complete from all sources. <sup>2</sup>11 variant carriers and 948 non-carriers had missing ethnicity information.

Based on these findings, we performed post-hoc analyses comparing variant-carriers and non-carriers in terms of: (i) mean Townsend index stratified by manifestation of a phenotype-of-interest; and, (ii) Townsend index breakdown by quintiles derived from the QResearch database^28,29^, comparing the frequency of phenotypes-of-interest. Variant-carriers had significantly higher levels of deprivation compared to non-carriers regardless of whether or not they manifested a phenotype-of-interest (−0.31 vs -1.07, p=0.0004 and -0.44 vs -1.4, p<2.2×10^−16^, respectively). Comparing the frequency of participants manifesting a phenotype, among variant-carriers it was similar in the least and most deprived quintiles (23% vs 24%), whereas among non-carriers, the least deprived had a lower frequency compared to the most deprived (17% vs 23%) (Supplemental Table 5).

Exploring the ethnicity distribution further, majority of *HTRA1* and *COL4A1* variant carriers (≥97%) were of self-reported White ethnicity. For *COL4A2*, however, 57% (190/336) of variant-carriers were of self-reported Black ethnicity, driven by 2 variants c.3448C>A and c.5068G>A present in 1.2% and 4.7% of Black participants, respectively. A further 14% of *COL4A2* variant-carriers were of mixed and other ethnic groups (Table 2).

**Table 2.** Variant-carriers in UK Biobank by gene and ethnic group.

| Gene | Self-reported ethnic group |  |  |  |  |  |  |
| --- | --- | --- | --- | --- | --- | --- | --- |
|  | All | White | Black | Asian | Mixed | Other | Unknown |
| *HTRA1* | 234 | 229<br>(98%) | 1<br>(0.4%) | - | - | 1<br>(0.4%) | 3<br>(1.3%) |
| *COL4A1* | 481 | 465<br>(97%) | 4<br>(0.8%) | 5<br>(1%) | 2<br>(0.4%) | 5<br>(1%) | - |
| *COL4A2* | 336 | 90<br>(27%) | 190<br>(57%) | 3<br>(0.9%) | 14<br>(4%) | 31<br>(10%) | 8<br>(2%) |
| Any gene-of-interest | 1,050 | 783 | 195 | 8 | 16 | 37 | 11 |

Legend: 'Other' ethnicity includes Chinese. There was one participant with a variant in both *HTRA1*
and *COL4A1*. There were no participants carrying variants in *TREX1* or *CTSA*.

### Assessing the proportion of variant carriers with phenotypes-of-interest

The proportion of variant carriers (N=1,050) with ≥1 phenotype-of-interest in the hospital inpatient admissions and/or death record data was: *HTRA1* 9% (21/234); *COL4A1* 20% (95/481); and *COL4A2* 4% (15/336) (Figure 2a). This proportion increased when we explored the smaller subset of the variant carriers for whom primary care data was also available (N = 484): *HTRA1* 55% (64/117); *COL4A1* 40% (93/236); and *COL4A2* 7% (9/132) (Figure 2b). Among variant carriers manifesting a phenotype-of-interest, stroke was not always the most common phenotype: *HTRA1* 13% to 52%, *COL4A1* 15% to 19% and *COL4A2* 93-100% (Figure 2a, Figure 2b).

**Figure 2.**
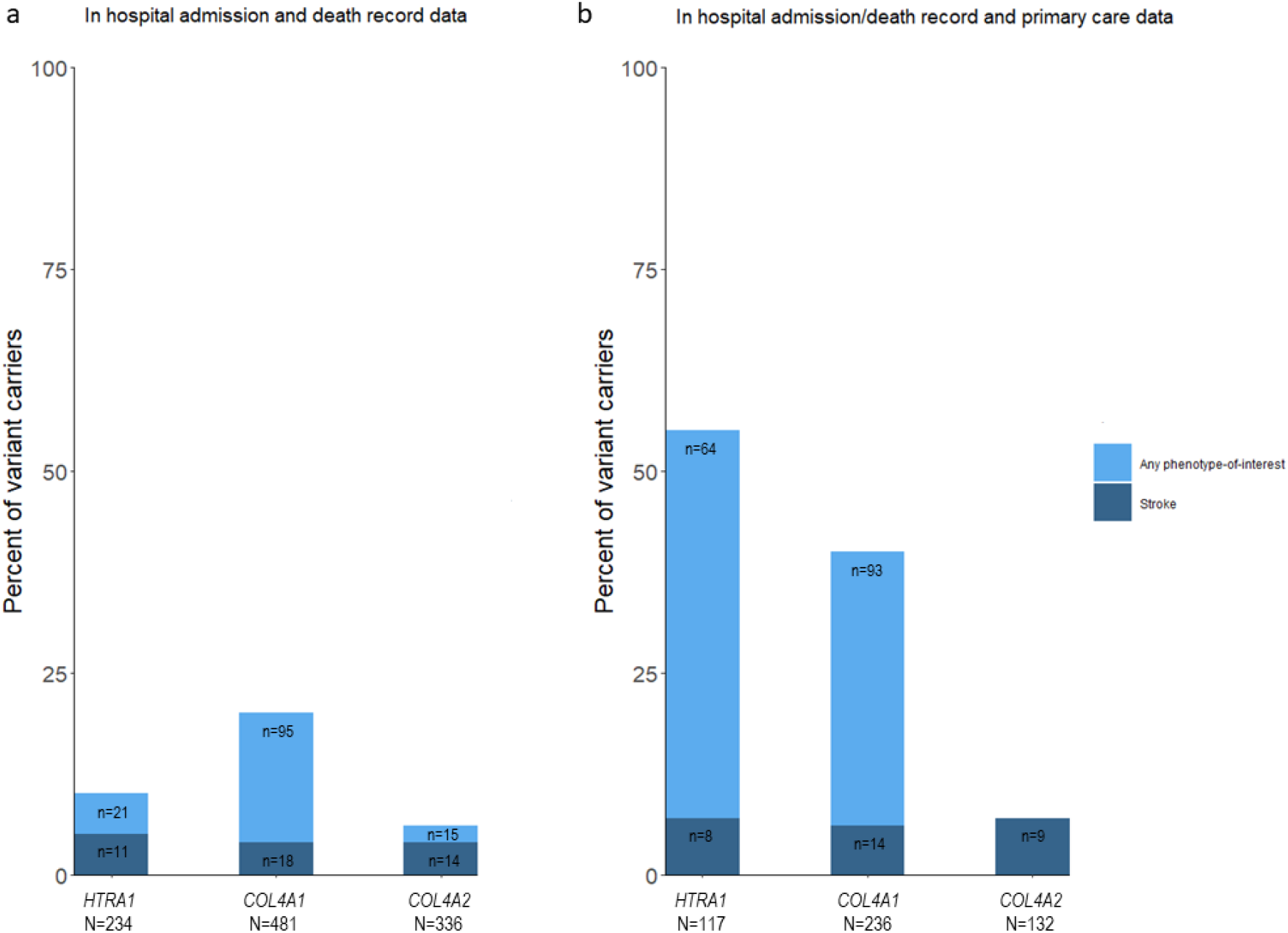
Proportion of variant carriers with a phenotype-of-interest. N=total number of variant-carriers; n=number of variant carriers with any phenotype-of-interest or stroke.

### Assessing whether variant carrier status is associated with phenotypes-of-interest

#### Association with phenotype-of-interest status

For phenotypes-of-interest in the hospital inpatient admissions and/or death record data, a higher proportion of *COL4A1* variant carriers compared to non-carriers had a *COL4A1*-related phenotype (20% in carriers vs 15% in non-carriers, p=0.01). We found no significant associations for *HTRA1* and *COL4A2*, and no significant associations for any gene in the secondary analyses also including primary care data. There was also no significant difference in the proportion of stroke cases seen between carriers and non-carriers for any of the genes.

#### Association with phenotype burden (phenotype score)

*COL4A1* carriers also had a greater phenotype score compared to non-carriers (OR=1.29, p=0.006). We found no significant associations for *HTRA1* and *COL4A2*, nor for any gene in the secondary analyses including primary care data. (Figure 3).

**Figure 3.**
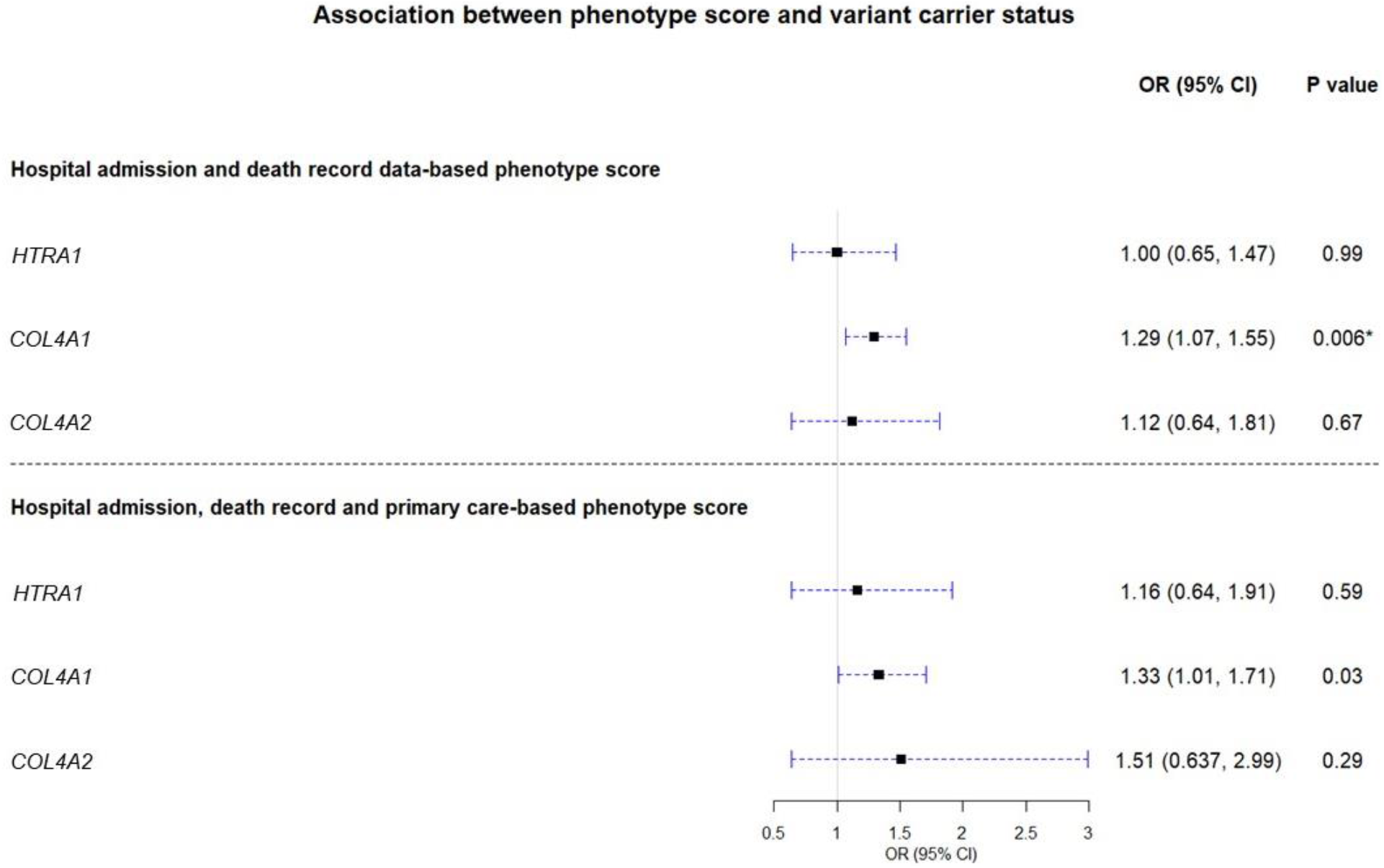
Association of variant carrier status with phenotype burden. OR=odds ratio; CI=confidence interval; *The association between *COL4A1* variant carrier status and phenotype score was significant.

The associations remained similar after adjusting the primary analyses for the presence of vascular risk factors (Supplemental Table 6). We found no significant interactions with sex (*HTRA1* p=0.41; *COL4A1* p=0.14; *COL4A2* p=0.49) and ethnicity (*HTRA1* p=0.60; *COL4A1* p=0.88; *COL4A2* p=0.19) for any gene.

Leave-one-out sensitivity analyses did not change the results significantly for *HTRA1* or *COL4A2* (Supplemental Figure 1). For *COL4A1*, removing cataract, migraine or stroke from the phenotype score rendered the association no longer significant, suggesting these phenotypes are important in driving the association seen in the primary analyses (Figure 4).

**Figure 4.**
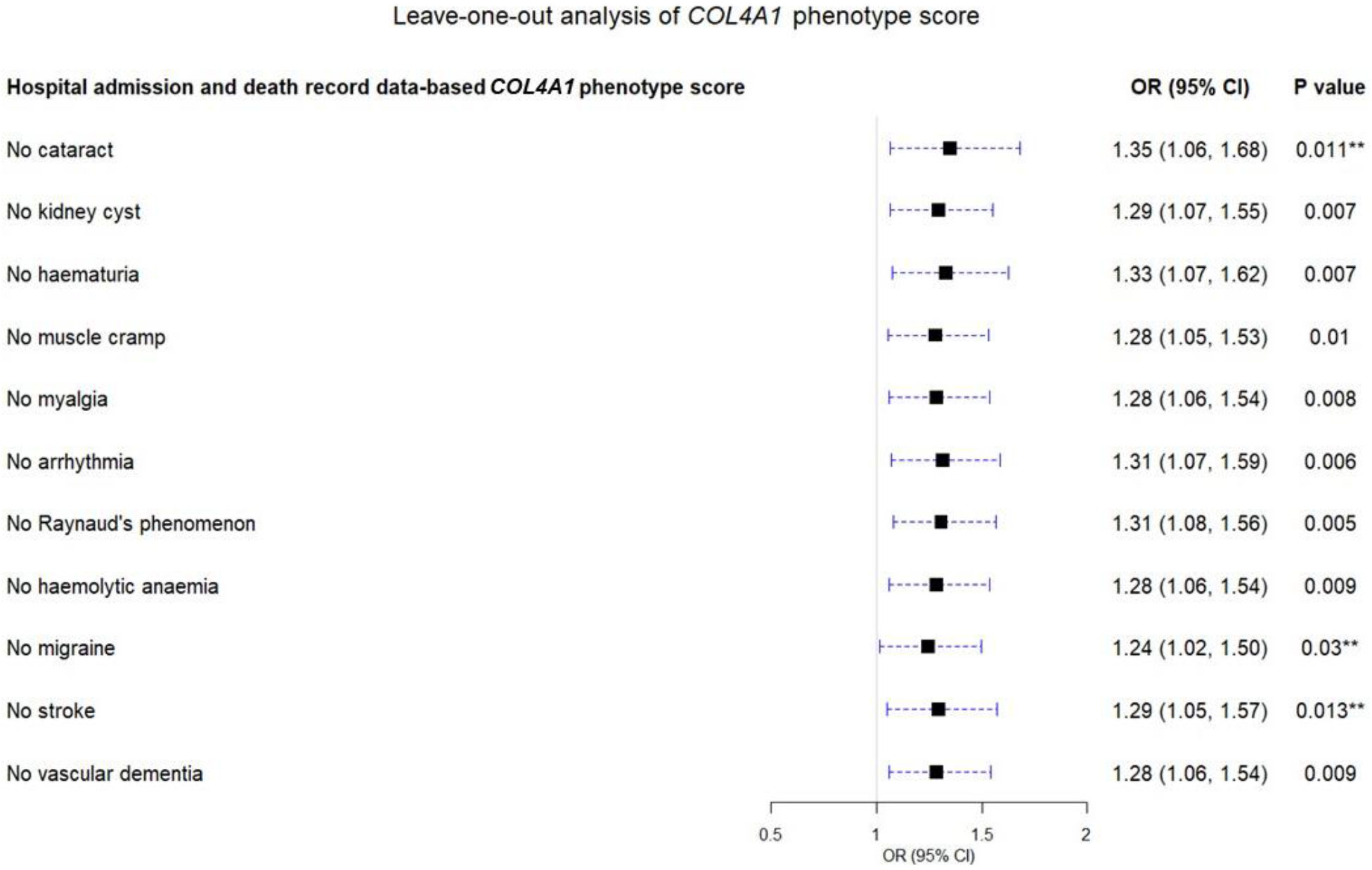
*COL4A1* leave-one-out analyses. OR=odds ratio; CI=confidence interval; **Association no longer significant.

## Discussion

We found that while 1:200 UKB participants carry a previously reported putative pathogenic rare variant in one of the five cSVD genes included in our study, only 4% to 20% of variant-carriers per gene had an associated phenotype recorded in their hospital admission/death records, and this rose moderately to 7% to 55% when also including primary care records. *COL4A1* variant carrier status was associated with having phenotypes-of-interest compared to non-carriers, but we did not see significant associations with expected phenotypes for other genes.

We are not aware of previous studies investigating these five genes in a population-based setting. There has however been a study of another monogenic cSVD gene, which demonstrated that ∼1:450 UKB participants carry a putative pathogenic (i.e., cysteine-altering) variant *NOTCH3*^19^. Among the few disease-based studies exploring rare variation in cSVD genes in apparently sporadic cases, one large study found that ∼1:70 lacunar stroke patients had a monogenic cause^13^. However, this study included only patients with stroke (one of several manifestations of cSVD), excluded already diagnosed monogenic cSVD cases and involved an overlapping but not identical set of genes with different definition of putative pathogenicity compared to our study, limiting direct comparisons.

We did not find any carriers of *CTSA* or *TREX1* putative pathogenic rare variants in UKB. Potential reasons include: (i) variants in these genes are extremely rare in general and/or rare in a population-based setting owing to their severe phenotype manifestations, this is particularly relevant for *TREX1* where the majority of variants were frameshifts which were not as highly represented in UKB WES data compared to single nucleotide variants^18^; (ii) the overall number of variants-of-interest in these genes was smaller and they are not present in UKB by chance.

We found a significantly higher level of deprivation among variant-carriers compared to non-carriers. Further post-hoc analyses suggested that this difference is not explained by variant-carriers manifesting a disease phenotype. Exploring the Townsend deprivation index distribution by quintiles suggested that carrying a putative pathogenic rare variant increases the chances of having a phenotype among the least deprived, but not among the most deprived participants. One possible explanation might be that participants in the most deprived group have an already increased risk due to environmental factors, whereas in the least deprived group, genetic variation plays a more important role. However, this was not the primary aim of our study and requires further research.

The majority of *COL4A2* carriers in UKB were of non-White self-reported ethnic group. The two genetic variants driving the frequency among non-White participants had previously been reported in the literature as causing intracerebral haemorrhage in persons of White Hispanic and African American background^30^. Earlier studies have focused mainly on investigating European populations, while case reports and series are often missing ethnicity information^6,13,31,32^, leaving monogenic cSVD and relevant genetic variation prevalence estimates among non-White ethnicities largely unknown^13,32^. One study which did investigate this in the Genome Aggregation Database among seven ethnic groups did not find a similar enrichment of *COL4A2* carriers among participants of Black and other ethnicities, although interestingly *COL4A1* pathogenic variants were most prevalent among Africans/African-Americans^31^. Furthermore, epidemiological studies have demonstrated racial and ethnic differences in cSVD manifestations and burden, which may have a genetic component^33^. These findings underline the importance of extending future genetic studies to include a broader range of ethnic groups.

Evidence from the existing literature summarised in a systematic review demonstrated a higher proportion of variant carriers manifesting a relevant phenotype compared to our study, with estimates of 59% for *COL4A2*, 75% for *COL4A1*, and 77% for *HTRA1*^6^. Considering the mean age of UKB participants is older than the mean age of individuals included in the systematic review^6^, it is unlikely this difference is explained by limited duration of follow-up in UKB and our analysis capturing participants who will go on to develop the phenotypes in the future. Our results suggesting lower frequency of phenotypic manifestations in carriers of variants associated with cSVD in a population-based setting are in keeping with findings from other monogenic disease investigations. Similar results have been shown for monogenic diabetes and CADASIL^19,34–36^, demonstrating ascertainment context is crucial when interpreting the consequences of monogenic variants. This has important implications for genetic screening and counselling in the clinical setting.

We identified a significant association for *COL4A1* between variant carrier status and phenotype score. Leave-one-out subgroup analyses indicated that migraine, cataract, and stroke contributed most to the association seen. We did not find significant associations between *HTRA1/COL4A2* and phenotypes previously attributed to putative pathogenic variants in those genes. This may suggest that these variants: (i) have reduced penetrance; (ii) have variable expressivity; (iii) are not all pathogenic despite previous reports; (iv) have previously been incorrectly associated with certain disease-phenotypes in the literature and OMIM; and/or (v) were not present in large enough numbers limiting statistical power.

Our variants-of-interest had evidence from the literature and ClinVar to support that they are pathogenic. Using Ensemble VEP to further investigate these variants generally corroborated this assumption - all but one had a moderate or high impact according to SnpEff and about two thirds were bioinformatically predicted to be deleterious and probably damaging to protein structure and function.

Our study has several strengths. By using all available sources of routinely collected health data, we were able to systematically capture the full range of phenotypes previously reported to be associated with monogenic cSVDs, both cerebral and extra-cerebral. This approach limits several of the biases which affect case-reports and series and is feasible among large numbers of participants. Our study is the first to explore rare variation in these five genes in a population-based setting, and as such provides valuable information on their frequency and spectrum of clinical consequences, supplementing existing knowledge derived mainly from case-reports and series. This in turn can inform clinicians of various specialties and clinical geneticist in selecting patients to test and when counselling variant carriers in the clinical setting.

There are also limitations which need to be considered. Firstly, we used routinely collected coded administrative health data to identify disease-phenotypes. While we made significant efforts to map the phenotypes-of-interest systematically and transparently to relevant disease codes, driven by clinically informed selection, these coded data are likely to identify some false-positive and miss true-positive cases. Also, it was more challenging to map some phenotypes than others, and some health conditions (e.g., alopecia, migraine or muscle cramps) are less likely to lead the person to seek medical help and hence be captured by the coded data. Secondly, the UKB population is highly likely affected by the ‘healthy volunteer bias’^16^, with clinically severely affected variant carriers less likely to enrol in the first place. Hence the variants identified among the UKB population may be those with lower overall penetrance, variable expressivity, and weaker evidence of pathogenicity. Thirdly, even when collapsing the variants across each gene, statistical power for detecting small and moderate variant-level genotype-phenotype association effects remained low. Fourth, routinely collected administrative disease codes did not provide data on cerebral radiological features of UKB participants, which is an important (and sometimes the only) manifestation of cSVD^4^.

Future studies will be required to extend these findings to other populations, aiming to better understand and minimise healthy volunteer biases and assessing a broader range of ages and socio-economic- and ethnic groups with even larger sample sizes. As methods of disease identification from routinely collected health data develop further, this will also allow more comprehensive and reliable capture of phenotypes-of-interest. Future work could also explore all rare variants in these genes, stratifying them by level of evidence for pathogenicity and using the richness of the routinely collected health data to undertake phenome-wide association studies.

In conclusion, putative pathogenic rare variants in five monogenic cSVD genes occur in the population at a frequency of 1:200, but only up to half of variant carriers have a relevant disease phenotype recorded in their linked health data. We could not replicate most previously reported gene-phenotype associations, suggesting lower penetrance rates, overestimated pathogenicity and/or limited statistical power. We also highlight the importance of considering the wider spectrum of phenotypic manifestations in cSVD.

## Supporting information

Supplemental methods: phenotype code selection protocol

Supplemental phenotype code list

## Data Availability

Original phenotype and genotype data is available from UK Biobank. ICD-10 and Read v2/3 code lists used for analysis are available in the Appendix (Supplemental code list).

## Supplemental data

Supplemental data includes six tables and one figure displaying additional variant information and results.

Supplemental methods: details the methodology used to map the phenotypes-of-interest to ICD-10 and Read v2/v3 codes.

Supplemental code list: contains the ICD-10 and Read v2/v3 code lists used to derive the phenotypes-of-interest for this study.

## Declaration of interests

The authors declare no competing interests.

## Acknowledgements

AF is funded by BHF award RE/18/5/34216. KR and AF are funded by Health Data Research UK Rutherford fellowship (MR/S004130/1). AT is funded by HDR-UK award HDR-9004 and HDR-9003. We would like to acknowledge Professor Martin Dichgans from the Institute for Stroke and Dementia Research in Munich Germany for his input at the earlier stages of conception of this work.

## Data and code availability

Original phenotype and genotype data is available from UK Biobank^16^. ICD-10 and Read v2/3 code lists used for analysis are available in the Appendix (Supplemental code list).

## Notes

### Competing Interest Statement

The authors have declared no competing interest.

### Author Declarations

UK Biobank https://www.ukbiobank.ac.uk/

