## Supplemental tables and figures for "Frequency and phenotype associations of rare variants in five monogenic cerebral small vessel disease genes in 200,000 UK Biobank participants with whole exome sequencing data"

**Supplemental Table 1.** Phenotypes previously attributed to our genes-of-interest.

| **Phenotype name** | **OMIM^1^ description** | **Description used for mapping to disease codes^2^** |
| --- | --- | --- |
| ***CTSA* phenotypes included in *CTSA* phenotype score (score range 0-7)** | | |
| Dry eyes | Dry eyes | Dry eyes |
| Dry mouth | Dry mouth | Dry mouth |
| Hypertension | Hypertension | Hypertension |
| Muscle cramps | Muscle cramps | Muscle cramps or spasms |
| Migraine | Migraine | Migraine, migraine with/without aura, complicated migraine |
| Stroke | Stroke | Any stroke subtype |
| Vascular dementia | Dementia, progressive | Vascular dementia |
| ***TREX1* phenotypes included in *TREX1* phenotype score (score range 0-8)** | | |
| Anaemia | Haemolytic anaemia | Normocytic/normochromic anaemia, non-autoimmune haemolytic anaemia |
| Liver disease | Micronodular cirrhosis of liver, abnormal liver enzymes | Liver disease, elevated liver enzymes, cirrhosis, portal hypertension, nodular regenerative hyperplasia, micro/macrovesicular steatosis, periportal inflammation, portal and bridging fibrosis, hepatic encephalopathy, liver failure, liver inflammation |
| Migraine | Migraine | Migraine, migraine with/without aura, complicated migraine |
| Nephropathy | Kidney glomerular dysfunction, proteinuria, haematuria | Elevated creatinine/urea, renal impairment, chronic renal failure, nephritic/nephrotic syndrome, proteinuria, haematuria, renal transplant |
| Raynaud’s phenomenon | Raynaud’s phenomenon | Raynaud’s phenomenon |
| Retinal vasculopathy | Retinal vasculopathy | Retinal vasculopathy, retinal vascular occlusions |
| Stroke | Stroke | Any stroke subtype |
| Vascular dementia | Dementia, progressive | Vascular dementia |
| ***HTRA1* phenotypes included in *HTRA1* phenotype score (score range 0-6)** | | |
| Back pain | Lumbago, lower back pain | Lower back pain |
| Degenerative spine disease | Lumbar disc herniation, spondylosis deformans | Spondylosis, sciatica, lumbar disc prolapse, disc herniation, disc displacement/degeneration, CT confirmed degenerative spine changes |
| Hair loss | Alopecia | Alopecia or baldness or hair loss with no further detail or diffuse hair loss/thinning or alopecia with male pattern baldness/frontal baldness |
| Migraine | Migraine | Migraine, migraine with/without aura, complicated migraine |
| Stroke | Stroke | Any stroke subtype |
| Vascular dementia | Dementia, progressive | Vascular dementia |
| ***COL4A1* phenotypes included in *COL4A1* phenotype score (score range 0-12)** | | |
| Anterior segment dysgenesis (ASD) | Anterior segment defects | Anterior segment dysgenesis, Axenfeld-Rieger anomaly, Peter’s anomaly, iris hypoplasia, posterior embryotoxon, corneal opacity, corneal clouding, congenital malformations of cornea/iris |
| Arrhythmia | Supraventricular arrhythmias | Supraventricular arrhythmia, atrial fibrillation |
| Cataract | Congenital cataract | Cataract, non-diabetic cataract |
| Haematuria | Haematuria | Haematuria |
| Haemolytic anaemia | Haemolytic anaemia | Non-autoimmune haemolytic anaemia |
| Kidney cysts | Renal cysts | Kidney cysts, cystic kidney |
| Migraine | Migraine | Migraine, migraine with/without aura, complicated migraine |
| Muscle cramp | Muscle cramps | Muscle cramps or spasms |
| Myalgia | Muscle cramps, elevated creatine kinase (CK) | Myalgia |
| Raynaud’s phenomenon (Raynaud’s) | Raynaud’s phenomenon | Raynaud’s phenomenon |
| Stroke | Stroke | Any stroke subtype |
| Vascular dementia | Dementia, progressive | Vascular dementia |
| ***COL4A2* phenotypes included in *COL4A2* phenotype score (score range 0-2)** | | |
| Stroke | Stroke | Any stroke subtype |
| Vascular dementia | Dementia, progressive | Vascular dementia |

^1^OMIM - Online Mendelian Inheritance in Man, <https://omim.org/>; ^2^informed by OMIM and systematic review (Rannikmäe *et al*. Stroke 2020;51:3007–3017).

**Supplemental Table 2.** Rare variants found among UK Biobank participants and their frequency.

| **Variants found in UKB** | **Resource** | **Number of variant carriers** | **Variant frequency (%)** |
| --- | --- | --- | --- |
| ***HTRA1*** | | | |
| c.754G>A^d^ | ClinVar & SysRev | 1 | 0.0005 |
| c.767T>C^d^ |  | 9 | 0.0045 |
| c.821G>A |  | 1 | 0.0005 |
| c.883G>A^d^ |  | 3 | 0.0015 |
| c.904C>T^ter^ |  | 4 | 0.002 |
| c.961G>A^d^ |  | 9 | 0.0045 |
| c.1108C>T^ter^ |  | 11 | 0.0055 |
| c.397C>G | SysRev | 2 | 0.001 |
| c.451C>A |  | 151 | 0.0755 |
| c.496C>T |  | 8 | 0.004 |
| c.517G>A |  | 2 | 0.001 |
| c.523G>A^d^ |  | 1 | 0.0005 |
| c.646G>A^d^ |  | 2 | 0.001 |
| c.958G>A^d^ |  | 24 | 0.012 |
| c.1348G>C |  | 6 | 0.003 |
| ***COL4A1*** | | | |
| c.2245G>A^d^ | ClinVar & SysRev | 2 | 0.001 |
| c.2159G>A^d^ |  | 1 | 0.0005 |
| c.1807C>T |  | 2 | 0.001 |
| c.3683G>T^d^ | ClinVar | 1 | 0.0005 |
| c.4717G>A^d^ | SysRev | 7 | 0.0035 |
| c.4213G>A^d^ |  | 1 | 0.0005 |
| c.3997G>A^d^ |  | 22 | 0.011 |
| c.3946C>G |  | 268* | 0.135 |
| c.3712C>T^d^ |  | 94 | 0.047 |
| c.3704A>G^d^ |  | 1 | 0.0005 |
| c.3671C>T^d^ |  | 33 | 0.0165 |
| c.3592G>A^d^ |  | 1 | 0.0005 |
| c.2641A>G |  | 11 | 0.0055 |
| c.1964G>A^d^ |  | 4 | 0.002 |
| c.1055C>T^d^ |  | 4 | 0.002 |
| c.196C>A |  | 28 | 0.014 |
| c.-2C>T |  | 1 | 0.0005 |
| ***COL4A2*** | | | |
| c.4147G>A^d^ | ClinVar & SysRev | 1 | 0.0005 |
| c.4357G>T^ter^ | ClinVar | 2 | 0.001 |
| c.1776+1G>A | SysRev | 1 | 0.0005 |
| c.3448C>A |  | 283* | 0.1415 |
| c.5068G>A |  | 50 | 0.025 |

*one participant was homozygous for the variant; ^ter^variants predicted to be nonsense mutations (early termination of protein/stop codon gained) by Variant Effect Predictor; ^d^variants predicted to be deleterious and probably damaging by Variant Effect Predictor;

**Supplemental Table 3.** Investigated and identified variants-of-interest by gene and resource.

ClinVar=ClinVar database; SysRev=systematic review; n=number of variants-of-interest identified among UKB participants; N=number of overall variants-of-interest.

| **Gene** | **Variants-of-interest** | | |
| --- | --- | --- | --- |
|  | **In ClinVar & SysRev**  **n/N** | **In ClinVar only**  **n/N** | **In SysRev only**  **n/N** |
| *CTSA* | 0/0 | 0/0 | 0/1 |
| *TREX1* | 0/0 | 0/0 | 0/10 |
| *HTRA1* | 7/19 | 0/6 | 8/30 |
| *COL4A1* | 3/35 | 1/36 | 13/94 |
| *COL4A2* | 1/6 | 1/6 | 3/10 |
| **Any gene** | **11/60** | **2/48** | **24/145** |

| **Summary of VEP output** | | | | | | | | | |
| --- | --- | --- | --- | --- | --- | --- | --- | --- | --- |
| **Gene** | **Both ClinVar & SysRev variants** | | | **SysRev variants** | | | **ClinVar variants** | | |
|  | **Variant consequence** | | | | | | | | |
|  | Intronic/  Untranslated region | Missense | Stop gained | Intronic/  Untranslated region | Missense | Stop gained | Intronic/  Untranslated region | Missense | Stop gained |
| *HTRA1* | - | 5 | 2 | - | 8 | - | - | - | - |
| *COL4A1* | - | 3 | - | 1 | 12 | - | - | 1 | - |
| *COL4A2* | - | 1 | - | 1 | 2 | - | - | - | 1 |
|  | **Variant impact/classification of severity (SNPEff)** | | | | | | | | |
|  | Low/ No information | Moderate | High | Low/ No information | Moderate | High | Low/ No information | Moderate | High |
| *HTRA1* | - | 5 | 2 | - | 8 | - | - | - | - |
| *COL4A1* | - | 3 | - | 0/1 | 12 | - | - | 1 | - |
| *COL4A2* | - | 1 | - | - | 2 | 1 | - | - | 1 |
|  | **Impact on protein function (SIFT)** | | | | | | | | |
|  | Tolerated | Deleterious | No information | Tolerated | Deleterious | No information | Tolerated | Deleterious | No information |
| *HTRA1* | 1 | 4 | 2 | 1 | 7 | - | - | - | - |
| *COL4A1* | - | 4 | - | 3 | 9 | 1 | - | 1 | - |
| *COL4A2* | - | 1 | - | 2 | - | 1 | - | - | 1 |
|  | **Impact on protein structure and function (PolyPhen)** | | | | | | | | |
|  | Possibly damaging | Probably Damaging | Benign/No information | Possibly damaging | Probably Damaging | Benign/No information | Possibly damaging | Probably Damaging | Benign/No information |
| *HTRA1* | 1 | 4 | 0/2 | 5 | 3 | - | - | - | - |
| *COL4A1* | 1 | 3 | - | 3 | 9 | 1 | - | 1 | - |
| *COL4A2* | - | 1 | - | 2 | - | 1 | - | - | 0/1 |

**Supplemental Table 4.** Summary of Ensembl Variant Effect Predictor (VEP) output.

ClinVar=ClinVar database; SysRev=systematic review;

**Supplemental Table 5.** Townsend deprivation index breakdown by quintiles**.**

| Townsend deprivation index |  | |
| --- | --- | --- |
| ALL PARTICIPANTS | | |
|  | Variant-carriers (N=1,048)^1^  % (N) | Non-carriers (N=198,013)^1^  % (n) |
| Quintile 1 | 26.2 (275) | 32.6 (64,582) |
| Quintile 2 | 20.4 (214) | 24.5 (48,437) |
| Quintile 3 | 18.5 (194) | 18 (35,615) |
| Quintile 4 | 17.4 (182) | 15 (29,781) |
| Quintile 5 | 17.5 (183) | 9.9 (19,598) |
| Participants with phenotypes-of-interest | | |
|  | Variant-carriers (N=226)  % (N) | Non-carriers (N=37,129)  % (N) |
| Quintile 1 | 27.4 (62) | 30.1 (11,183) |
| Quintile 2 | 21.2 (48) | 24 (8,917) |
| Quintile 3 | 15.5 (35) | 17.9 (6,646) |
| Quintile 4 | 16.8 (38) | 15.8 (5,872) |
| Quintile 5 | 19 (43) | 12.1 (4,511) |
| Participants without phenotypes-of-interest | | |
|  | Variant-carriers (N=822)  % (N) | Non-carriers (N=160,884)  % (N) |
| Quintile 1 | 25.9 (213) | 33.2 (53,399) |
| Quintile 2 | 20.2 (166) | 24.6 (39,520) |
| Quintile 3 | 19.3 (159) | 18 (28,969) |
| Quintile 4 | 17.5 (144) | 14.9 (23,909) |
| Quintile 5 | 17 (140) | 9.4 (15,087) |
| DERIVED frequency of phenotypes-of-interest | | |
|  | Variant-carriers (N=1,048)  % with phenotype-of-interest | Non-carriers (N=198,013)  % with phenotype-of-interest |
| Quintile 1 | 23% | 17% |
| Quintile 2 | 22% | 18% |
| Quintile 3 | 18% | 19% |
| Quintile 4 | 21% | 20% |
| Quintile 5 | 24% | 23% |

Quintile 1=least deprived; Quintile 5=most deprived; N=number of participants. ^1^Information about the Townsend index was missing for 2 variant carriers and 250 non-carriers.

**Supplemental Table 6.** Association of variant carrier status with phenotype burden adjusted for presence of vascular risk factors

|  | **OR (95% CI)** | **P-value** |
| --- | --- | --- |
| **Hospital admission/death record data derived phenotype scores** | | |
| *HTRA1* | 0.97 (0.62, 1.42) | 0.91 |
| *COL4A1* | 1.29 (1.06, 1.54) | 0.008* |
| *COL4A2* | 1.15 (0.65, 1.86) | 0.61 |

OR=odds ratio; CI=confidence interval; *significant association; Vascular risk factors: body mass index (<30 versus ≥30), smoking status (non-smoker versus current/previous smoker), alcohol consumption (≤twice a week versus > twice a week), hypertension and diabetes (present versus absent).

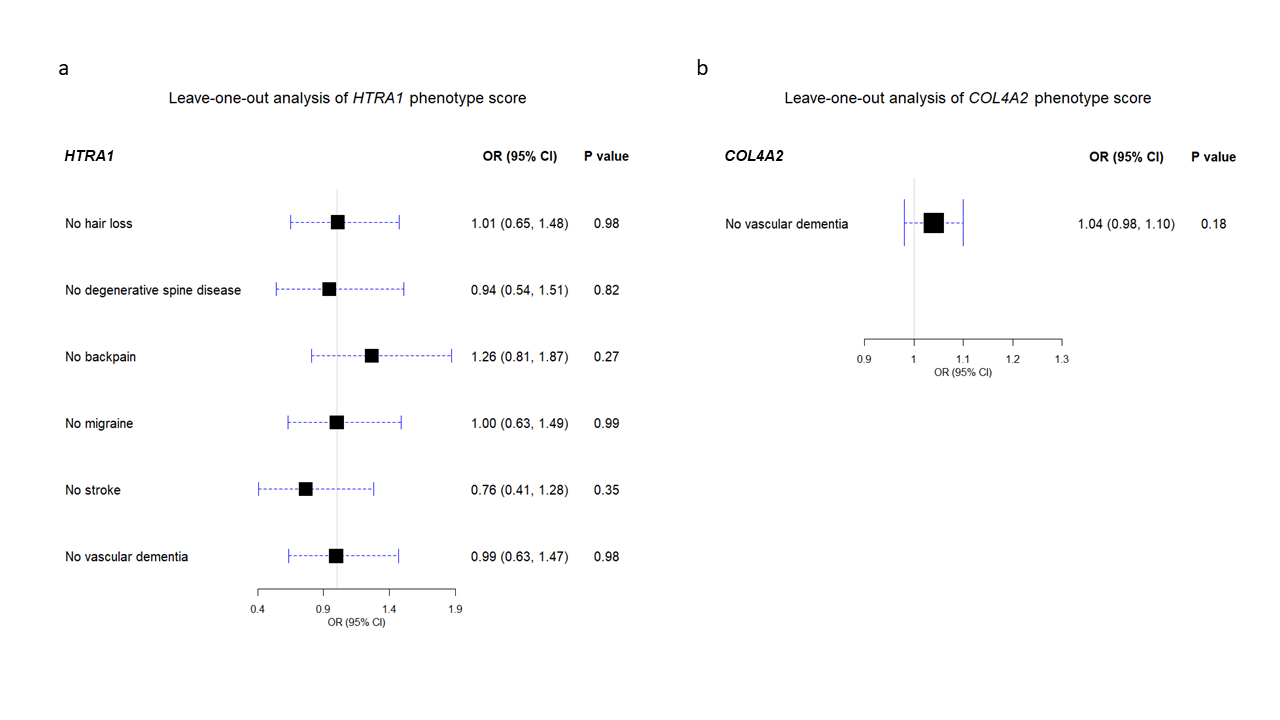

**Supplemental Figure 1**. *HTRA1* and *COL4A2* Leave-one-out analysis using hospital admission/death record data.

OR=odds ratio; CI=confidence interval.
