## Supplemental methods: phenotype code selection protocol for "Frequency and phenotype associations of rare variants in five monogenic cerebral small vessel disease genes in 200,000 UK Biobank participants with whole exome sequencing data"

**Steps and rules for creating phenotype code lists**

**Table of contents:**

1. Resources used

1a. Resources used for looking up code lists for our phenotypes of interest, and mapping codes from different coding systems to each other.

1b. Resources used for finding pre-existing code lists for our phenotypes of interest.

2. Process of creating relevant code lists for our phenotypes of interest, and mapping codes from different coding systems to each other

3. Process for selecting codes to include in the final list

4. Challenges and caveats

Step 2. Process of creating relevant code lists for our phenotypes of interest, and mapping codes from different coding systems to each other.

Step 3. Process for selecting codes to include in the final list.

5. Our lessons and conclusions

**1. Resources used**

**1a. Resources used for looking up code lists for our phenotypes of interest, and mapping codes from different coding systems to each other**

a) International Statistical Classification of Diseases and Related Health Problems 10th Revision from <https://icd.who.int/browse10/2019/en>, and UK Biobank Data-Field 41270, to search for relevant ICD-10 codes.
b) UK Biobank Data-Field 20002 to search for self-reported non-cancer illness codes.
c) From <http://www.surginet.org.uk/informatics/opcs.php> website and UK Biobank Data-Coding 240 for relevant OPCS-4 codes.
d) UK Biobank resource 592 “Clinical coding classification systems and maps”. This resource includes excel spreadsheets with full code lists and their descriptions (including Read v2, Read v3, ICD-9, ICD-10 and OPCS-4), and provides information on how to map from Read v2 and Read v3 to other clinical coding systems. It also specifies that the accuracy of code lists, definitions and maps should be verified by specialists as part of any analysis undertaken on these data.

**1b. Resources used for finding pre-existing code lists for our phenotypes of interest**

e)	CALIBER  <https://caliberresearch.org/portal/codelists>

f)	UK Biobank Category 42 Algorithmically-defined outcomes, Health-related outcomes

**2. Process of creating relevant code lists for our phenotypes of interest, and mapping codes from different coding systems to each other**

At this stage, codes were assessed for relevance to the phenotype of interest and there was a low threshold to extract all potentially relevant codes. The steps below were undertaken by a senior medical student (Miss Sophie Thrippleton) and a junior doctor (Dr David Henshall), both with significant prior experience of working with clinical codes in the research and/or clinical setting.

1) We used the ICD-10 coding system as the starting point and foundation for code selection. First, we reviewed the ICD-10 coding trees from resources ‘a’ and ‘d’ above, and identified relevant ICD-10 codes.
2) We used resource ‘d’ to map relevant ICD-10 codes to Read v3 codes.
3) We used resource ‘d’ to map relevant Read v3 codes to Read v2 codes.
4) We reviewed mapped Read v2 and Read v3 codes to ensure they made sense from the clinical perspective.
5) We conducted an additional manual search to identify any additional Read v2 and Read v3 codes, which may have been missed by the mapping done using resource ‘d’.
6) We compared our resulting code lists with already existing code lists for phenotypes where this was available from resource ‘e’ or ‘f’, and added any additional codes not already included.
7) We reviewed the list of self-reported non-cancer illness codes (resource ‘b’) and identified relevant codes.
8) We reviewed the list of OPCS-4 codes (resource ‘c’) and identified relevant codes.

**3. PROCESS FOR Selecting codes to include in the final list**

At this stage, all relevant codes from all coding systems were reviewed by the whole team. Decisions about inclusion/exclusion were made jointly following the protocol below (Figure 1). In case of uncertainty or disagreement, final decisions were made by Dr Kristiina Rannikmäe (clinical academic neurology consultant). When selecting renal disease codes, we also consulted with a clinical academic nephrology consultant, Dr Bryan Conway.

**Figure 1. Process for selecting codes to include in the final list.**

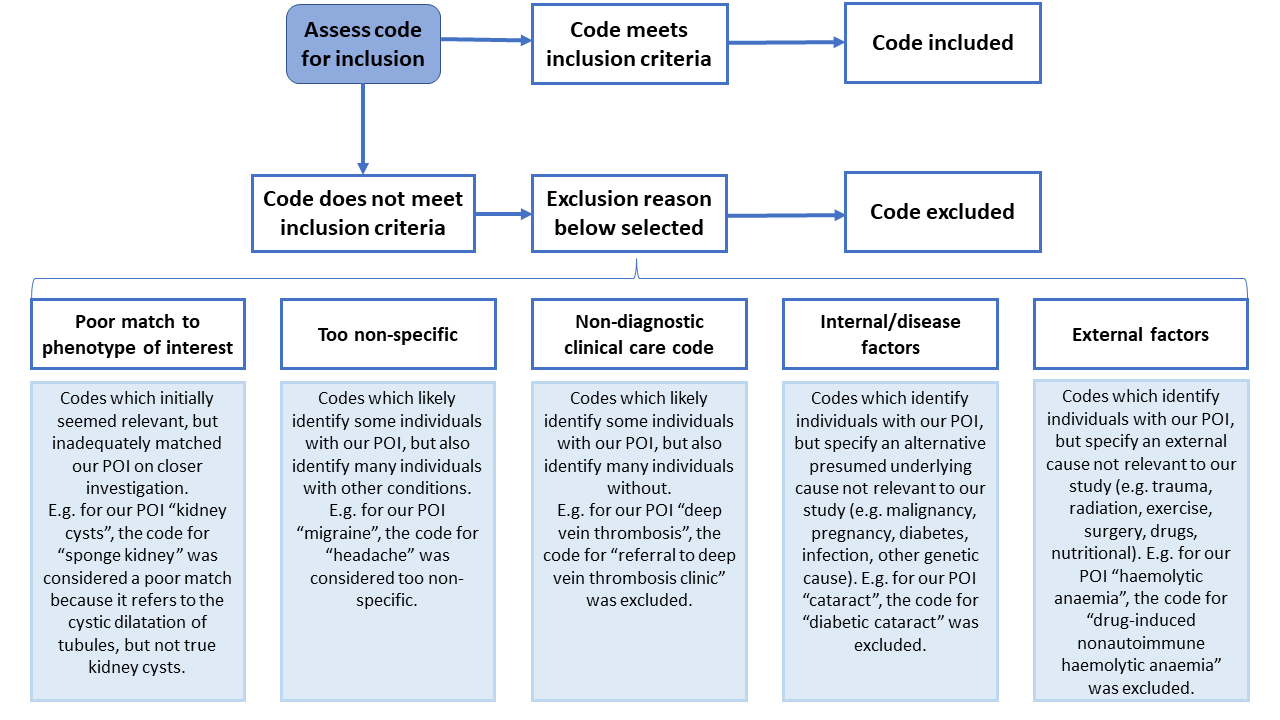

POI = phenotype of interest

**4. challenges and caveats**

**Step 2: Process of creating relevant code lists for our phenotypes of interest, and mapping codes from different coding systems to each other:**

- ICD-10 codes displayed in <https://icd.who.int/browse10/2019/en> and in UK Biobank Data-Field 41270 varied slightly which is why we used both resources to compile a comprehensive list (e.g. ‘M54.5’ for ‘Low back pain’ has a further subdivision to denote specific areas of lower back pain in the UK Biobank Data-Field only: ‘M54.50’ for ‘Low back pain, multiple sites in spine’, ‘M54.54’ for ‘Low back pain – Thoracic’ etc.).
- In Read v2 and Read v3 coding systems, one code can be linked to ≥1 description of the same clinical concept. For example, a ‘Myocardial Infarction’ may be referred to as a ‘Heart Attack’. Over time, some synonyms have been added that have a different meaning to the preferred term. Therefore, we only considered the preferred terms in our code selection and mapping processes.
- For a small number of Read v2 codes, the mapping files in resource ‘d’ did not have clinical descriptions. If this was the case, the mapped Read v3 clinical description was used instead.
- The mapping files in UK Biobank Data-Field 41270 contain errors and clinical knowledge is needed to screen the results (e.g. the mapping file maps ICD-10 code ‘Q619’ for ‘Cystic kidney disease, unspecified’ to Read v3 code ‘XM00P’ for ‘Primary microcephaly’).
- We conducted an additional manual search to identify any additional Read v2 and Read v3 codes, which may have been missed by the mapping done using resource ‘d’ (mapping files in UK Biobank Data-Field 41270). This step turned out to be very important as it identified a significant proportion of additional codes for some of our phenotypes of interest (e.g. for ‘kidney cyst’, 46 relevant codes were identified from the mapping file, while a manual search identified an additional 28 codes for this phenotype).

**Step 3. Process for selecting codes to include in the final list:**

- The main challenge we faced was clearly defining our phenotypes of interest. This was particularly difficult for symptoms, where a broad range of underlying pathologies could be associated with the symptom (e.g. haematuria).
- Exclusion reason ‘too non-specific’ was at times hard to apply consistently. In reality, the threshold was set on a phenotype-by-phenotype basis. These decisions are subjective judgements and factor in disease-related knowledge, how refined the phenotype of interest is, number of codes available and numbers of individuals with certain codes.
- Exclusion reason ‘Internal/disease factors’ also caused some challenges. For example, where definite causality between the factor and phenotype is uncertain (e.g., struvite kidney stones and infection), or where an infection can be a trigger for another intrinsic disease process (e.g., IgA nephropathy).

**5. OUR LESSONS AND CONCLUSIONS:**

- Clinical insight is needed to select appropriate code lists from available resources.
- Considering the many challenges and caveats, documenting the process and transparency is very important for replication.
- Detailing the rationale for exclusion of any potentially relevant codes may allow more meaningful sensitivity analyses and allows the option of re-defining cases & controls by filtering in/out certain codes as appropriate.
- Selecting a ‘good’ code list is also dependent on how the codes are used in real world practice e.g., if general practitioners only use ‘high level’ and ‘unspecified’ codes, then code lists which carefully pick disease subtype codes will not yield desired results.
